# The development and usability testing of digital knowledge translation tools for parents of children with acute otitis media

**DOI:** 10.1101/2021.06.29.21259431

**Authors:** Anne Le, Lisa Hartling, Shannon D. Scott

## Abstract

Acute otitis media (AOM) is the most common bacterial ear infection affecting up to 80% of children before the age of three. Despite the common occurrence of the illness and the wide range of material available at clinics and online, parents are not always aware of these resources and they are often difficult to understand. We worked with parents to develop and assess the usability of a whiteboard animation video and interactive infographic for AOM in children. Parents rated the tools highly across all usability items, suggesting that creative tools developed using multi-method development processes can be useful, relevant, understandable, and will be used by the intended audience.

Following the completion of the English-language products, our team culturally adapted the tools for the Pakistan context and evaluated the usability of these adapted tools. During usability testing, parents indicated that they felt the tools were useful, demonstrating that culturally adapted version of knowledge translations tools are effective in ensuring that parents could understand complex health information.

## Introduction

Acute otitis media (AOM) is a common infection of the middle ear that affects approximately 80% of children before the age of three. Caused by viruses, such as the respiratory syncytial virus (RSV), or bacteria, such as Streptococcus pneumoniae, symptoms of AOM are non-specific and can include irritability, ear pain, and fever [1-3]. Uncertainty about the cause and symptoms of AOM often results in difficulty with at-home management, diagnosis, and treatment. Furthermore, significant discrepancies are found in practice, with some professionals treating with antibiotics, while others allowing infections to resolve spontaneously [4]. The ambiguity surrounding the diagnoses and treatment of AOM in children have been found to be associated with parental distress. Studies have revealed that parents have limited and inaccurate knowledge about AOM, suggesting that more effective information sources and mediums are required to educate parents [5, 6].

Storytelling is one of the oldest methods of communication that spans across all cultures and generations [7]. Previous studies have found that narratives have potential to convey complex health information from providers to patients and have been associated with increased patient health outcomes [8, 9].

As co-directors of Translating Emergency Knowledge for Kids (TREKK), our research programs have developed and evaluated arts-based KT tools, demonstrating their use as effective sources of consumer-friendly, evidence-based information for parents. Combining parental narratives, results from systematic literature reviews, and best practice guidelines, we have developed an innovative method to equip parents with information and strategies to manage common childhood conditions (e.g., gastroenteritis, chronic pain, and croup).

The purpose of this study was to work with parents to develop and assess the usability of a whiteboard animation video and interactive infographic for AOM in children.

## Methods

This multi-method study employed patient engagement to develop, refine, and evaluate a whiteboard animation video and interactive infographic for pediatric acute otitis media. Research ethics approval was obtained from the University of Alberta Health Research Ethics Board (Edmonton, AB) [Pro00062904], Izaak Walton Killam (IWK) Research Ethics Board (Halifax, NS), and University of Saskatchewan Research Ethics Board (Saskatoon, SK). Operational approvals were obtained from individual emergency departments to conduct usability testing.

### Compilation of Parents’ Narratives

Parental narratives were informed through 16 semi-structured qualitative interviews (**Appendix A**) and a systematic review. Parents of children who presented to the Stollery Children’s Hospital (Edmonton, Canada) with AOM were invited to participate in interviews conducted by a project coordinator trained in qualitative methodology. Parents were asked about their experiences with pediatric AOM. Concurrently, a systematic review was conducted to synthesize current evidence about experiences and information needs of parents and caregivers managing acute otitis media. Results from the systematic review and qualitative interviews are published elsewhere [5].

### Intervention Development

Information gathered from the systematic review and interviews were used to develop the video script and infographic skeleton. Information from the TRanslating Emergency Knowledge for Kids (TREKK) Bottom Line Recommendations (BLR) for acute otitis media was included in the script and skeleton, which were then shared with a creative team to develop the e-tool prototypes [10]. Information pertinent to management of AOM were embedded within the storyline of the video, which depicted a couple struggling to manage their child’s ear pain (**Appendix B**). Likewise, information on symptoms, treatment options, and when to seek emergency care was included in the infographic (**Appendix C**).

### Revisions

Iterative processes were used to develop the AOM video and infographic. Different stakeholder groups, including parents, health care professionals (HCPs), researchers, and the study team participated in several rounds of revisions. HCPs were asked to comment on the quality and accuracy of information and evidence, length of the tool, aesthetics, usefulness, and perceived value. Parents from our Pediatric Parent Advisory Group (P-PAG) were asked to provide feedback on the length, stylistic elements, and highlight areas not addressed in the tools. The P-PAG meets once a month and are asked to participate in tool development several times a year [11]. Likewise, our research team also holds weekly meetings to discuss the development of our tools.

### Video

Over the course of 18-months, researchers worked with parents, HCPs, and the creative team to refine the tool for usability testing. Six versions of the video were developed before the AOM video was deemed ready for usability testing. The usability testing video was 3 minutes and 5 seconds in length and featured a family of three whose child presents with symptoms of AOM. The storyline starts with a family (father, mother, child) at home preparing for bed when Sam develops a slight fever and ear pain. The mom, serving as the narrator, describes the parents’ concern for their child. The parents then take the child to their family physician, resulting in an AOM diagnosis. The mom highlights information about AOM, as provided by their doctor, including a description of the condition and symptoms, what medications can be given, how to manage the condition at home, and when to seek healthcare and/or emergency care. Equipped with this information, the parents then decide to manage Sam at home. Unfortunately, their child’s symptoms worsen the next day, prompting a visit to the ED, where they are prescribed antibiotics. The video further reiterates the course of actions that parents should take when their child has symptoms of AOM. As the narration plays, images and graphics appear on screen to further highlight the information being provided. The family doctor also prescribes antibiotics and explains that they should only be used if symptoms become worse (i.e., persisting pain and/or fluid coming from the child’s ear). The parents then go home and Sam’s AOM resolves shortly after. The characters were designed to be realistic, while certain features, such as sex, age, and race were kept ambiguous. The style of the video was whiteboard animation, which is characterized by having a hand “draw” and “write” in images and texts that appear across the screen.

The video was later entered into the Canadian Institutes of Health Research (CIHR) Institute of Human Development, Child and Youth Health (IHDCYH) 2019 Talks Video Competition and received special commendations.

### Infographic

The AOM interactive infographic used a style that had been previously developed and tested for our pediatric procedural pain interactive infographic. This style which is unique to our tools underwent dozens of different iterations and took two years to develop. In order to ensure that our tools were consistent across all conditions, we used the same style, design elements, animations, and characters across all conditions in our suite of interactive infographic tools.

Our interactive infographic tools are developed to be similar to a webpage, such that they allow users to scroll through the information and explore at their own pace. Unlike videos or e-Books, such tools give parents the control to view what they need based on their needs. Information provided in our AOM interactive infographic was similar to information provided in our AOM video and consisted of seven different panels. These included: What is Acute Otitis Media?, Common Signs of AOM, Here’s What You Can Do, What You Can Do At Home, Antibiotics, Additional Resources, and Contact Us. Our KT Tool Development Cycle can be viewed in **Figure 1**.

**Figure 1.**
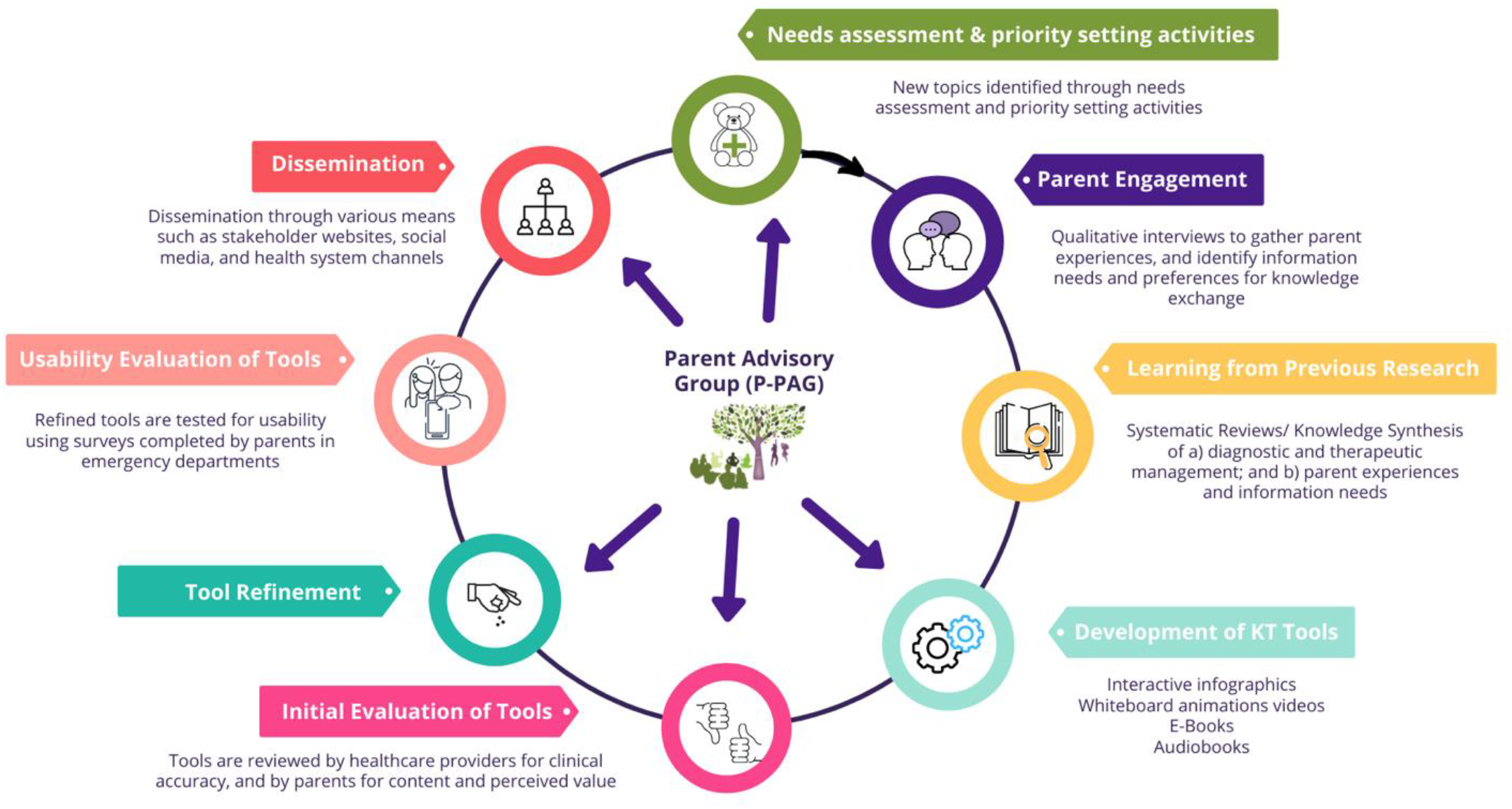
ARCHE-ECHO KT Tool Development Cycle

### Surveys

Parents were recruited to participate in an electronic, usability survey (**Appendix D**) in rural and urban ED waiting rooms in Alberta, Saskatchewan, and Nova Scotia. These included the Cobequid Community Health Centre, Hants Community Hospital, and Colchester Community Health Centre in Nova Scotia, the Royal University Hospital in Saskatchewan, and Leduc Community Hospital in Alberta. Surveys were informed by a systematic review of over 180 usability evaluations and comprised of 9, 5-point Likert items assessing: 1) usefulness, 2) aesthetics, 3) length, 4) relevance, and 5) future use [12]. Parents were also asked to provide their positive and negative opinions of the tool via two free text boxes. Members of the study team approached parents in the ED to determine interest and study eligibility. Study team members were available in the ED to provide technical assistance and answer questions as parents were completing the surveys.

### Data Analysis

Data were cleaned and analyzed using SPSS v.24. Descriptive statistics and measures of central tendency were generated for demographic questions. Likert answers were given a corresponding numerical score, from 5 to 1, with 5 being “Strongly Agree” and 1 being “Strongly Disagree” [13-14]. T-tests were conducted to determine whether there were any significant differences between the mean scores for both tools. Open-ended survey data were analyzed thematically. A summary of the results was then shared with the creative team to inform the development of the final versions of the tools.

## Results

58 parents participated in the study. Of those, 28 reviewed the whiteboard animation video, while 30 viewed the interactive infographic. Table 1 provides demographics of the sample.

**Table 1.** Demographic characteristics of parents who assessed the usability of the whiteboard animation video and infographic (N=58)

| Characteristic | n (%) |
| --- | --- |
| Sex |  |
| Female | 42 (72.4) |
| Male | 16 (27.6) |
| Age |  |
| 20-29 years | 7 (12.1) |
| 30-39 years | 20 (34.5) |
| 40-49 years | 24 (41.4) |
| 50-59 years | 6 (10.3) |
| 60 years and older | 1 (1.7) |
| Marital Status |  |
| Married/Partnered | 48 (82.8) |
| Single/Separated/Divorced/Widowed | 10 (17.2) |
| Education |  |
| Some high school | 2 (3.4) |
| High school diploma | 12 (20.7) |
| Some post-secondary | 7 (12.1) |
| Post-secondary certificate/diploma | 17 (29.3) |
| Post-secondary degree | 15 (25.9) |
| Graduate degree | 4 (6.9) |
| Household Income |  |
| Less than \$25,000 | 3 (5.2) |
| \$25,000-\$49,000 | 14 (24.1) |
| \$50,000-\$74,000 | 12 (20.7) |
| \$75,000-\$99,000 | 12 (20.7) |
| \$100,000-\$149,000 | 8 (13.8) |
| \$150,000 and over | 6 (10.3) |
| Prefer not to answer | 3 (5.2) |
| Number of Children in the Family |  |
| 1 | 12 (20.7) |
| 2 | 19 (32.8) |
| 3 | 19 (32.8) |
| 4 | 3 (5.2) |
| 5+ | 5 (8.5) |

**Table 2.**
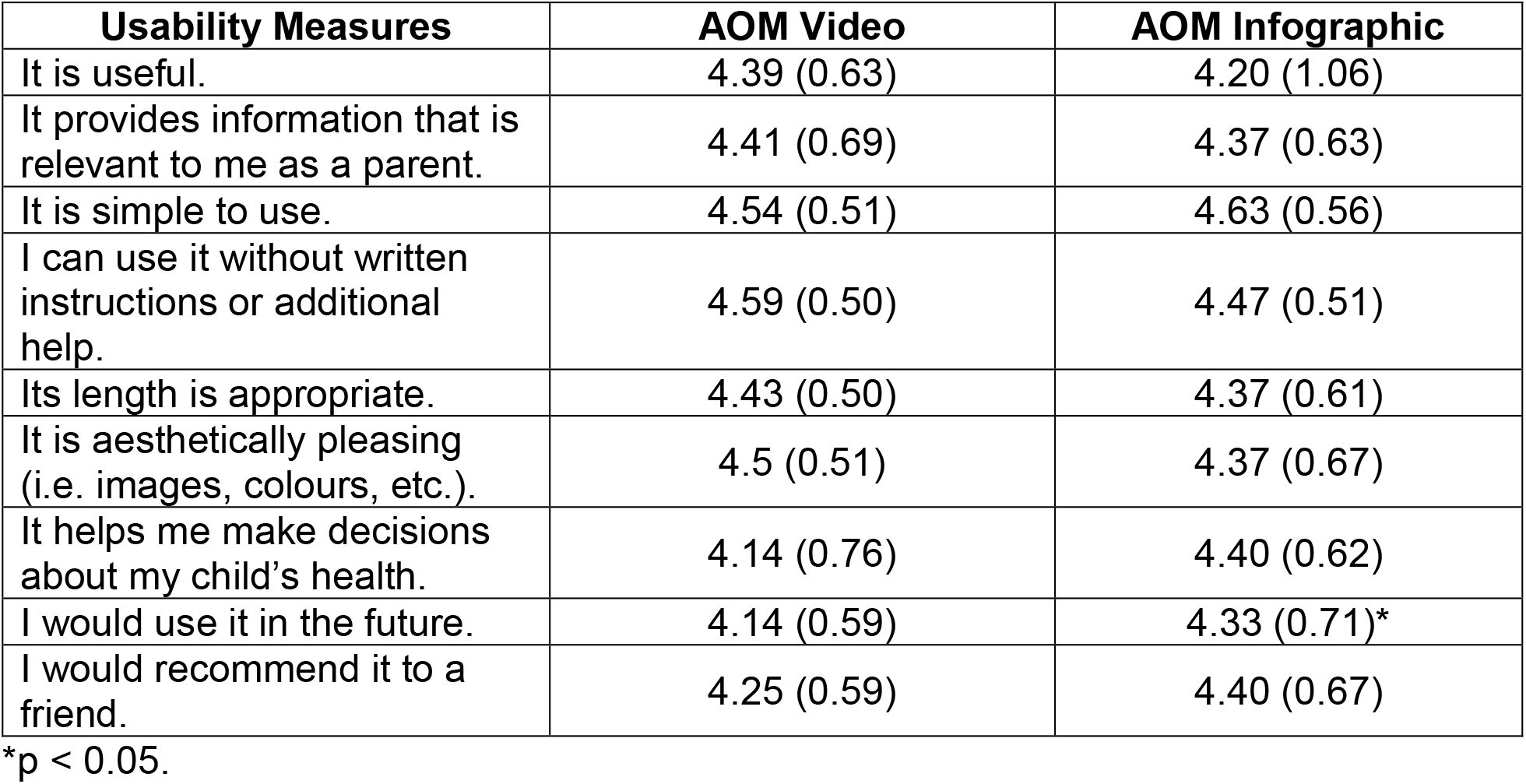
Means (SD) of participant responses to the usability survey

In general, parents rated the AOM whiteboard animation video positively, resulting in mean scores of at least 4.10 out of 5.00 on all questions. All parents strongly agreed or agreed that the tool was simple to use (4.53) and that it could be used without written instructions or help (4.59). Likewise, parents felt that the length of the video was appropriate (4.43) and that the tool was aesthetically pleasing (4.50). Parents gave high scores when asked if they found the tool useful (4.39) and if the tool was relevant to them as parents (4.41). When asked about potential future usage of the tool, scores were 4.14 for both “I would use it in the future” and “it will help me make decisions about my child’s health”. Finally, when asked if parents would recommend the tool to a friend, parents gave a score of 4.25.

Likewise, parents liked the interactive infographic, providing mean scores of 4.20 out of 5.00 on all questions. The highest score was given when we asked parents whether the tool was simple to use (4.63) while the lowest score was given for “it is useful” (4.20). Means were all 4.37 for relevancy, length, and aesthetics. Similarly, parents gave identical scores for “it helps me make decisions about my child’s health” and “I would recommend it to a friend” (4.40). Finally, scores for future use and using the tool without additional help were 4.33 and 4.47, respectively. When comparing the two tools, only future use scores were significantly different (p<0.05). Table 3 provides a breakdown of the means in usability items for each tool.

In the open text boxes, parents described the AOM video as “very helpful”, “clear”, and “easy to follow”. One parent wrote, “I wish more parents could view this, as ear infections seem to be what takes a lot of kids to the doctor unnecessarily!” while another discussed the tool as a potential learning tool for older children, “Voice is clear, easy to understand, provided information I did not know. I found the tool very useful and would help the older children understand what their diagnosis is and would possibly help them feel better to know what is going on inside them”.

Parents described the AOM infographic as “useful”, “clear”, and felt that it provided “quick and easy access to information”. Parents also stated that the infographic was appealing to look at while providing helpful information. Due to the positive scores and feedback from usability testing with parents, very few changes were made to the AOM tools. As such, the final iterations for usability testing were released for public consumption.

Following the completion of the English-language products, our team culturally adapted these tools for use in Pakistan. This project was funded by WCHRI [15]. Outputs and images of the Pakistani tools can be found on page 13 and on the **ECHO KT website** (www.echokt.ca).

## Conclusions

We developed and evaluated the usability of two innovative, arts-based, digital tools for parents with children with acute otitis media. Using comprehensive methods, our AOM tools were developed over the course of two years and involved several important steps: (1) synthesis of evidence from the literature; (2) collection of our own evidence through interviews with parents who have had children with AOM; (3) content outline development; (4) tool design and development; (5) several rounds of revisions and modifications with stakeholders and the study team, and (6) usability testing with our end-users, parents. High scores across all usability items for both tools suggest that using multi-method development processes can result in tools that are useful, relevant, understandable, and will be used by the intended audience.

Our tools are easily accessible and can be viewed across a broad range of devices, including desktop computers, mobile phones, and tablet devices. Providing a means for quick and convenient access ensures that parents can obtain information about how to manage their child’s condition at the tip of their fingers. Such resources may not only help parents save time but may also alleviate the burden on the health system, such that it provides information for parents to make evidence-based informed decisions to manage their child’s condition at home.

### The tools can be found here: http://www.echokt.ca/tools/ear-infection/

Note: Our KT tools are assessed for alignment with current, best-available evidence every two years. If recommendations have changed, appropriate modifications are made to our tools to ensure that they are up-to-date [16].

## Data Availability

Data referred to in the manuscript are not available for access.

## Other Outputs from this Project

## Outputs from the Pakistan AOM Project

## Appendices

### Appendix A – Interview Guide

Parents will be interviewed to understand their experience having a child with otitis media. Semi-structured interviews will be conducted with parents in order to get their “narrative” or experiences. The following questions will be used to guide these interviews. Being true to semi-structured interview techniques, interview questions will start broad and then move to the more specific.

1. Tell me about your experience having your child experience otitis media.
2. Tell me about your child that was ill. How old is your child? How was your child ill? Has your child previously had otitis media?
3. How did you feel during this experience?
4. What did you do to manage symptoms of otitis media? (any techniques you used, for example, giving Tylenol, talking with family/friends, etc.)
5. What strategies were put in place by health care professionals to help your child? (for example, giving/prescribing antibiotics). Did they ask you to do anything?
6. How did your child manage the experience? How did you feel about the outcome of this situation?
7. If presented with the same situation again, would you do anything differently? If so, please tell me.

### Appendix B – Video Images

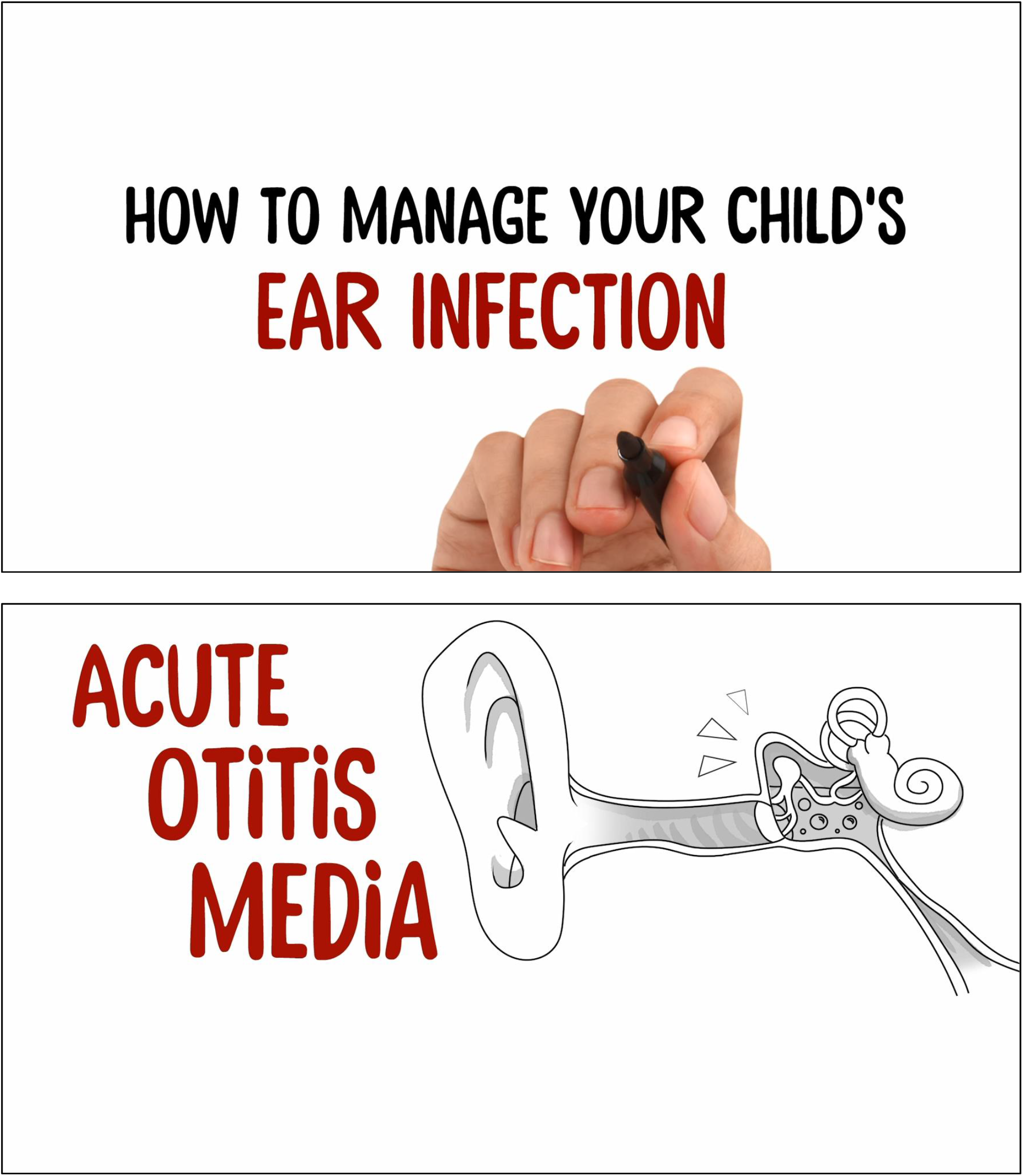
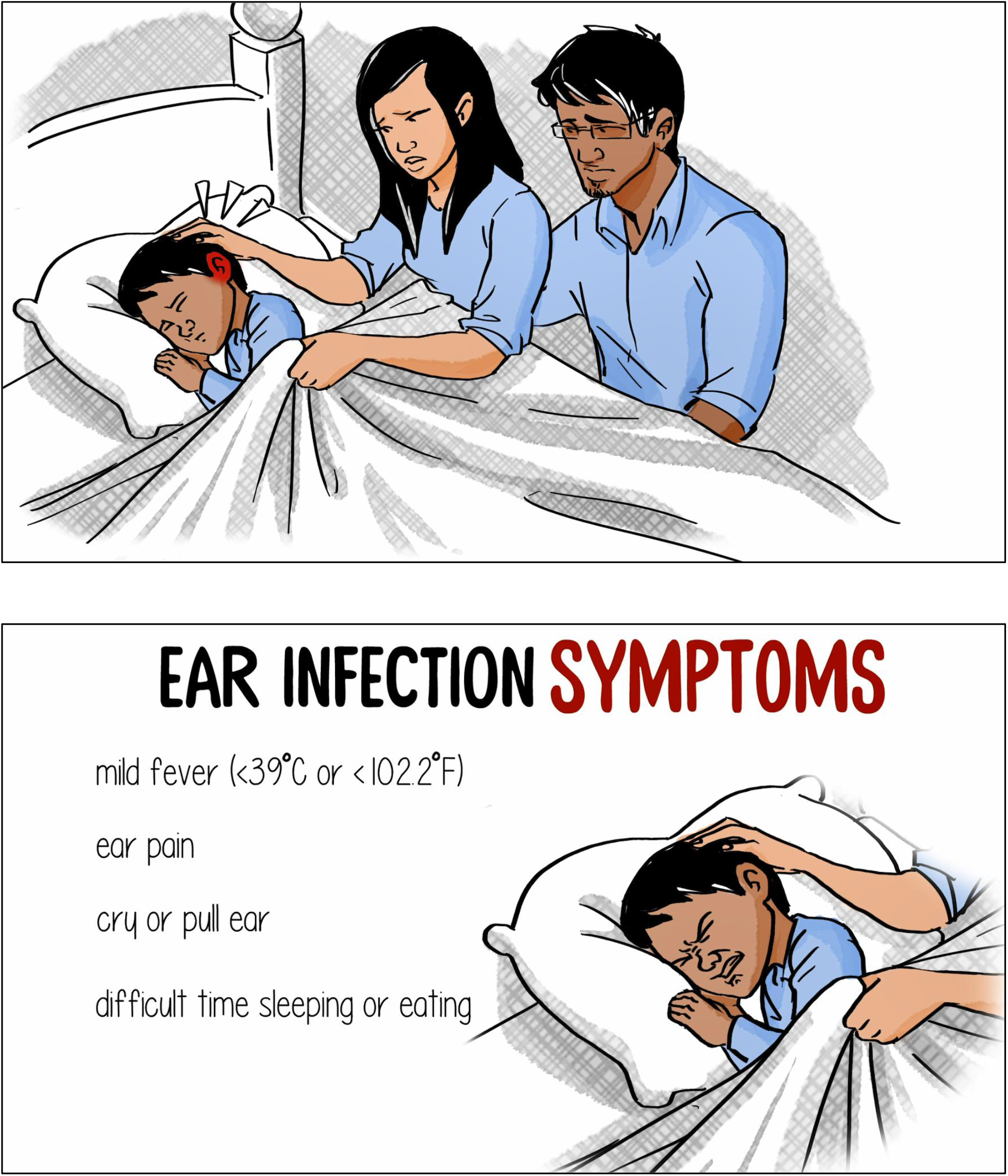
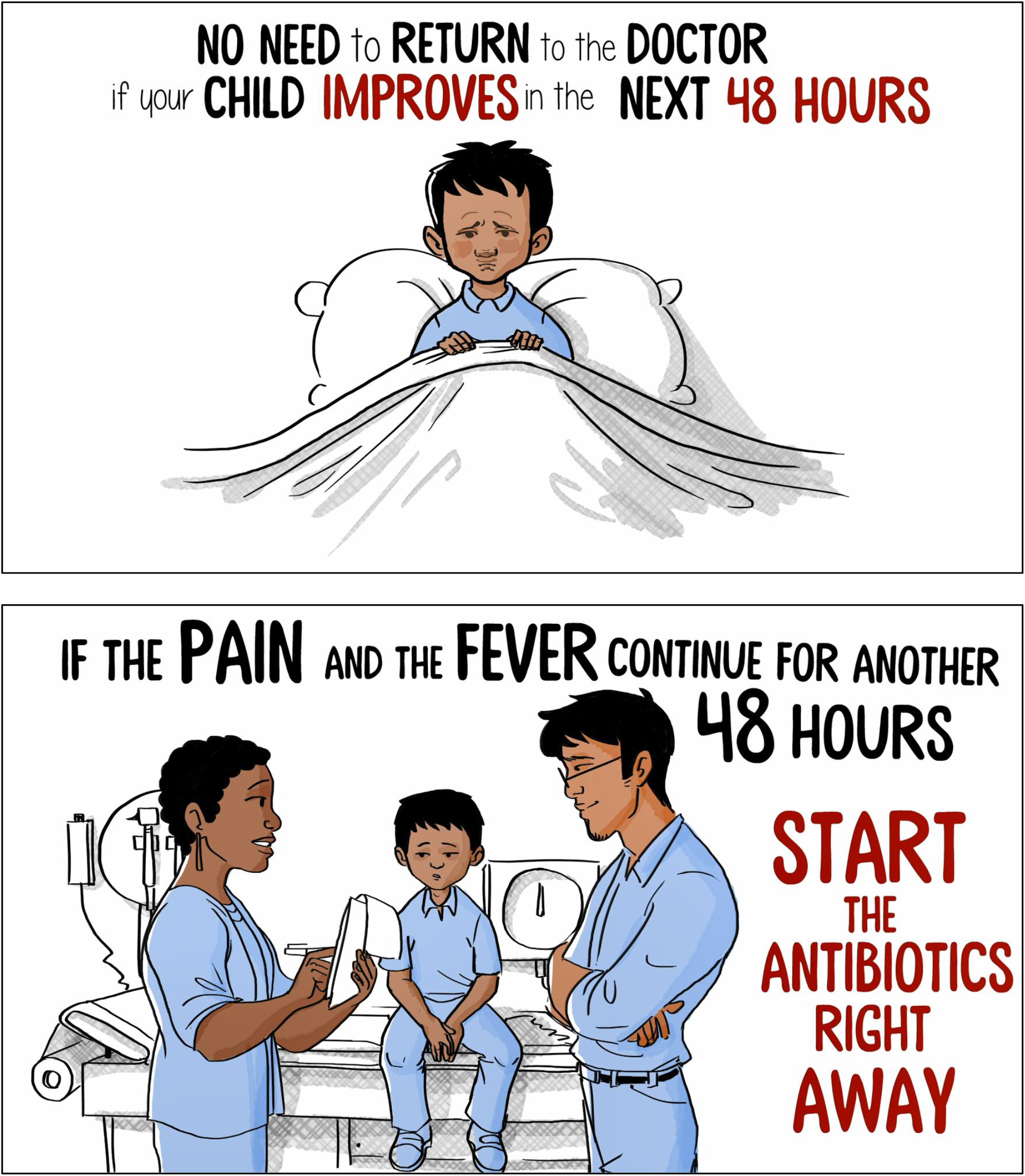

### Appendix C – Infographic Images

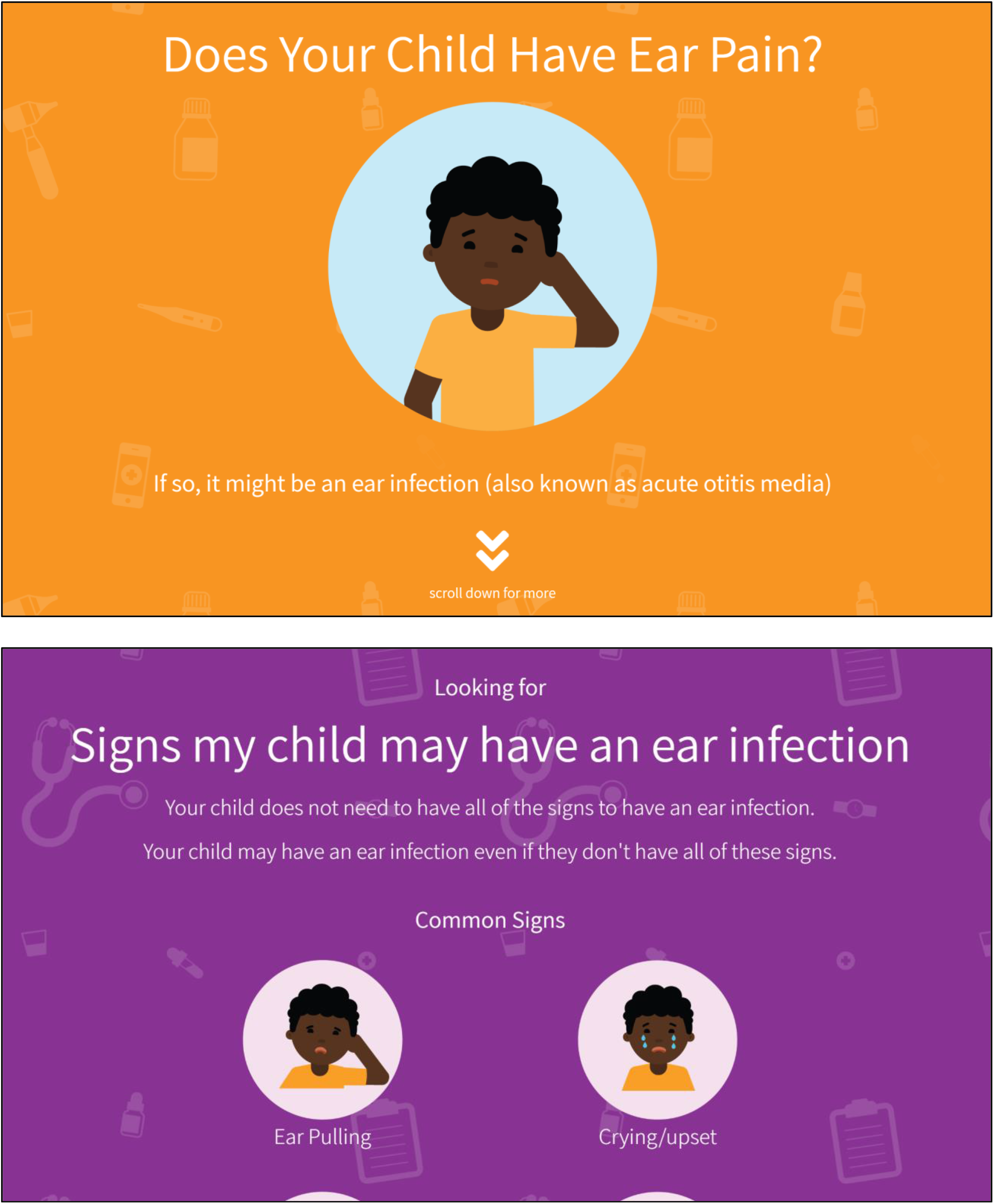
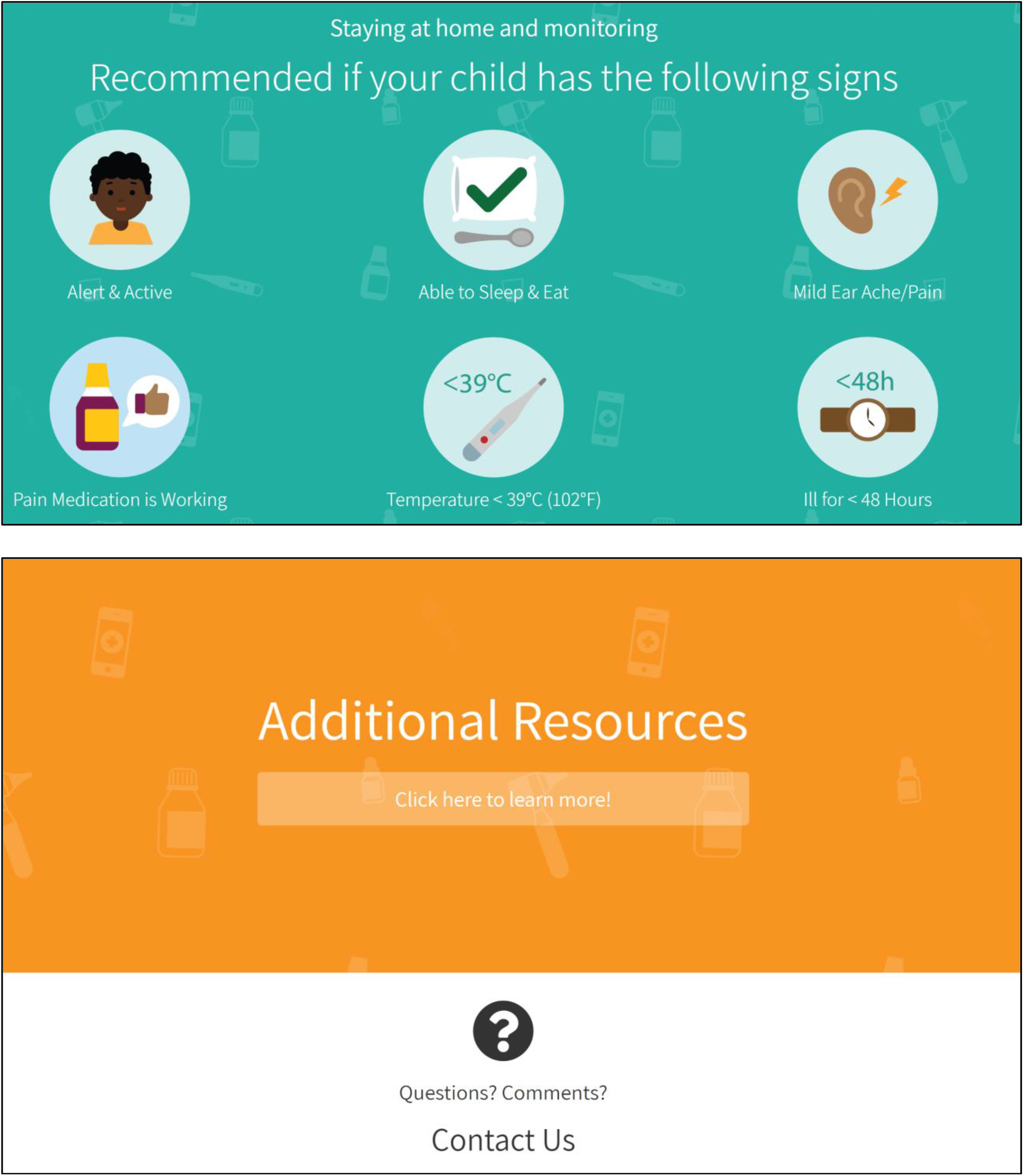

### Appendix D – Usability Survey

SECTION 1: Demographics

1) What is your gender?
  □ Male
  □ Female
3) What is your Age?
  □ Less than 20 years old
  □ 20-30 years
  □ 31-40 years
  □ 41-50 years
  □ 51 years and older
4) What is your Marital Status?
  □ Married
  □ Single
5) What is your gross annual household income?
  □ Less than $25,000
  □ $25,000-$49,999
  □ □ $50,000-$74,999
  □ □ $75,000-$99,999
  □ □ $100,000-$149,999
  □ □ $150,000 and over
6) What is your highest level of education?
  □ Some high school
  □ High school diploma
  □ Some post-secondary
  □ Post-secondary certificate/diploma
  □ Post-secondary degree
  □ Graduate degree
  □ Other
7) How many children do you have? ——
8) How old are your children? —————

SECTION 2: Assessment of attributes of the arts-based, digital tools

***\*participant is randomized to view 1 of 2 digital tools then automatically directed to the survey***

1. It is useful. [5-point Likert Scale]
2. It provides information that is relevant to me as a parent. [5-point Likert Scale]
3. It is simple to use. [5-point Likert Scale]
4. I can use it without written instructions or additional help. [5-point Likert Scale]
5. Its length is appropriate. [5-point Likert Scale]
6. It is aesthetically pleasing (i.e., images, colours, etc.). [5-point Likert Scale]
7. It helps me to make decisions about my child’s health. [5-point Likert Scale]
8. I would use it in the future. [5-point Likert Scale]
9. I would recommend it to a friend. [5-point Likert Scale]
10. List the most negative aspects: [open text]
11. List the most positive aspects: [open text]

### Appendix E – Project Timeline

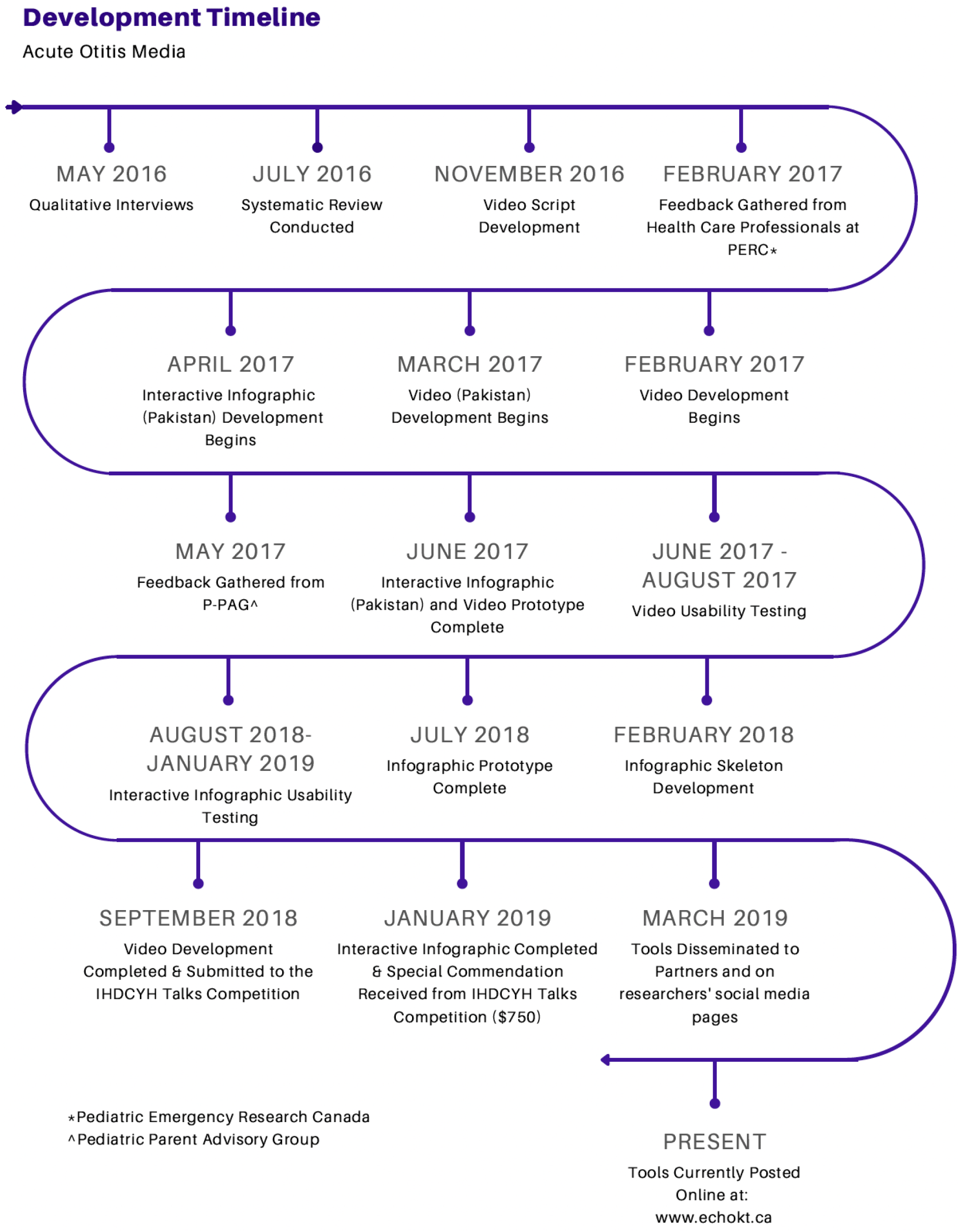

## Notes

### Competing Interest Statement

The authors have declared no competing interest.

### Funding Statement

This study was funded by the Networks of Centres of Excellence of Canada and the Women and Children's Health Research Institute

### Author Declarations

University of Alberta Research Ethics Board (REB)

